# Feasibility of generalised DKI approach for brain glioma grading

**DOI:** 10.1101/2020.09.23.20195719

**Authors:** E.L. Pogosbekian, I.N. Pronin, N.E. Zakharova, A.I. Batalov, A.M. Turkin, T.A. Konakova, I.I. Maximov

## Abstract

**Purpose:** An accurate differentiation of brain glioma grade constitutes an important clinical issue. Powerful non-invasive approach based on diffusion MRI has already demonstrated its feasibility in glioma grade stratification. However, the conventional diffusion tensor (DTI) and kurtosis imaging (DKI) demonstrated moderate sensitivity and performance in glioma grading. In the present work, we apply generalised DKI (gDKI) approach in order to assess its diagnostic accuracy and potential application in glioma grading.

**Methods:** Diffusion scalar metrics were obtained from 50 patients with different glioma grades confirmed by histological tests following biopsy or surgery. All patients were divided into two groups with low- and high-grade gliomas as II grade versus III and IV grades, respectively. For a comparison, trained radiologists segmented the brain tissue into three regions with solid tumour, oedema, and normal appearing white matter. For each region we estimated the conventional and gDKI metrics including DTI maps.

**Results:** We found high correlations between DKI and gDKI metrics in high-grade glioma. Further, gDKI metrics enabled introduction of a complementary measure for glioma differentiation based on correlations between the conventional and generalised approaches. Both conventional and generalised DKI metrics showed quantitative maps of tumour heterogeneity and oedema behaviour. gDKI approach demonstrated largely similar sensitivity and specificity in low-high glioma differentiation as in the case of conventional DKI method.

**Conclusion:** The generalised diffusion kurtosis imaging enables differentiation of low and high grade gliomas at the same level as the conventional DKI. Additionally, gDKI exhibited higher tissue contrast between tumour and healthy tissue and, thus, may contribute as a complementary source of information on tumour heterogeneity.

## Introduction

Primary brain glioma is a highly widespread type of intra-axial brain tumours seen approximately in one fourth diagnosed tumour cases [1]. Classification of the brain tumours is performed in accordance with grades introduced by the World Health Organisation (WHO) [2] which combine histopathological and molecular features into integrated tumour characterisation. As a result, glioma grades are ordered from I up to IV grade related to tumour’s aggressiveness and malignancy. Both the tumour’s diagnosis and its grade can be reliably confirmed by a histopathologic analysis of the tumour’s tissue obtained from an invasive procedure such as biopsy or surgery. However, non-invasive assessments of the tumour’s malignancy are necessary for clinical treatment, surgery planning and survival rate estimations. In particular, this applies to low-grade tumour cases where such assessments contribute to treatment efficacy and, as a result, improve quality of life for patients. Magnetic resonance imaging (MRI) offers a wide spectrum of tissue visualisation approaches for clinical purposes allowing one to detect and localise brain’s abnormalities with a certain degree of specificity. Nevertheless, conventional MRI techniques such as structural T_1_/T_2_-weighted imaging with or without contrast agents or magnetic resonance spectroscopy demonstrate limited sensitivity and specificity for brain glioma differentiation [3], [4].

Diffusion weighted imaging (DWI) has exhibited its superiority as a non-invasive brain imaging technique and has been applied in a plethora of clinical settings [5], [6], [7]. Diffusion tensor imaging (DTI) [8], the most often used DWI approach, has also been utilised for tumour assessment by many research groups [9], [10], [11]. In turn, diffusion kurtosis imaging (DKI) [12] is a powerful extension of the conventional DTI technique enabling estimation of the degree of non-Gaussian diffusion in the brain tissue by estimating higher order cumulant of the diffusion signal expansion [13]. The deviation of water molecule diffusion from free diffusion behaviour or Gaussian distribution is caused by complex organisation of the brain tissue, where cellular and neurite barriers alter the probability distribution of water diffusion. This phenomenon is particularly important for the brain tumour detection and assessment due to increased tissue complexity resulting from the high cancer cell proliferation rate, increased tumour tissue vascularisation, presence of oedema as well as necrosis. DKI is extensively used in brain imaging [14], [15], [16], in particular, for tumour differentiation [17], [18], [19], [20], [21]. Despite the fact that DKI demonstrated quite promising results in the case of glioma grading, the kurtosis scalar metrics such as mean kurtosis, still lack the accurate and reliable glioma grading in the case of both low- (i.e. glioma-I and II) and high- (glioma-III and IV) grade gliomas [18], [22], [23].

In the present study, we adapted the generalised kurtosis approach (GK) [24] for glioma differentiation in order to evaluate as well as to add a complementary information and, thus, increase the accuracy of glioma grading. An advantage of the third order term of cumulant expansion of the diffusion signal has been demonstrated for differentiation of low- and high-grade gliomas together with an application of the conventional kurtosis (CK) approach. In order to emphasise the advantage of the DKI imaging for glioma grading, we concurrently applied both DKI and DTI metrics derived from the CK and GK approaches. Hence, the purpose of the present study was to investigate the value of the generalised DKI metrics for the evaluation of glioma grading.

## Materials and Methods

### Patients

50 adult patients (mean age: 44 ± 13 years, minimal age: 24 years, maximal age: 70 years; male/female ratio: 30/20) initially diagnosed with glioma were recruited at our Institution and examined with MRI prior to biopsy, surgery, radiation or chemotherapy treatments. The diagnosis of glioma and WHO grade [25] were confirmed by histological and immunohistochemical examinations for each subject following MRI examination.

The study included 19 patients with glioma grade II, 8 patients with glioma grade III, and 23 patients with glioma grade IV. The group of patients with glioma-II consisted of 6 subjects with oligoastrocytoma, 1 subject with oligodendroglioma, 1 subject with hemistocytic astrocytoma, 11 subjects with diffuse astrocytoma. The group of patients with glioma-III consisted of 7 subjects with anaplastic astrocytoma, 1 subject with anaplastic oligoastrocytoma. The group of patients with glioma-IV consisted of the 22 subjects with glioblastoma multiform and 1 subject with gliosarkoma.

### Imaging

All patients underwent MRI examination with a 3T GE scanner (SignaHDxt) using a 8-channel phased-array head coil. The imaging protocol included precontrast T_2_-weighted FLAIR and diffusion-weighted sequences, followed by postcontrast T_1_-weighted imaging with a gadolinium based contrast agent. The parameters specific to anatomical imaging sequences were the following: T_1_-weighted: FOV = 240 mm^2^, TE/TR = 3500/8800 ms, resolution 1 mm^3^; T_2_-weighted FLAIR: FOV = 240 mm^2^, TE/TR = 120/9500 ms, resolution 1×1×5 mm^3^; diffusion-weighted imaging: FOV = 240 mm^2^, TE/TR = 102.8/10000 ms, resolution 3 mm^3^, *b*-values = 0, 1000, and 2500 s/mm^2^ with 60 encoding diffusion directions, directions were non-coplanar uniformly distributed over the unit sphere. The diffusion imaging was obtained using a single-shot echo echo planar imaging sequence.

### Image Analysis

Prior to estimation of scalar diffusion metrics, the raw diffusion data were corrected for image distortions in accordance with optimised pipeline [26], i.e. for noise [27] and Gibbs-ringing distortions [28], eddy-current distortions including bulk head motion artefacts were corrected using *eddy* utility from FSL [29], [30], [31]. Conventional DKI metrics such as mean, axial, radial kurtosis, and kurtosis anisotropy (MK, AK, RK and KA, respectively) were estimated using an iterative weighted least squares approach [32] and supplied by the complementary DTI measures known as fraction anisotropy (FA), mean diffusivity (MD), axial diffusivity (AD), and radial diffusivity (RD). The conventional DKI metrics were obtained from the second order cumulant expansion in the following form [12]:

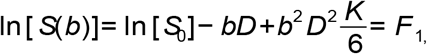

where *b* is the diffusion weighting, *D* is the diffusion coefficient, *S*_0_ is the signal without the diffusion weightings, and *K* is the diffusion kurtosis. The righthand expression is defined as *F*_1_. The generalised DKI parameters were estimated using in-house MATLAB scripts (The MathWorks, Natick, MA USA) including the third order terms [24]:

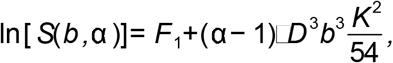

where α is the adjusting variable. In the case of α = 1, the last equation leads to the conventional DKI expression *F*_1_. In the present work, we used an optimised α value at 2/7 [24]. Subsequently, the estimated diffusion scalar maps for both cases of conventional and generalised kurtosis were aligned and interpolated to T_1_-weighted anatomical image for each patient using the cubic spline interpolation algorithm and affine transformation. Two independent radiologists manually delineated three regions of interest for each subject: tumour, oedema and normal appearing white matter (NAWM) using 3 MRI contrasts: T_1_-weighted, T_2_-weighted, and MD map estimated from CK approach. Examples of images for low- and high-grade gliomas with the 3 aforementioned regions of interest are presented in Figures 1 and 2, respectively.

**Figure 1.**
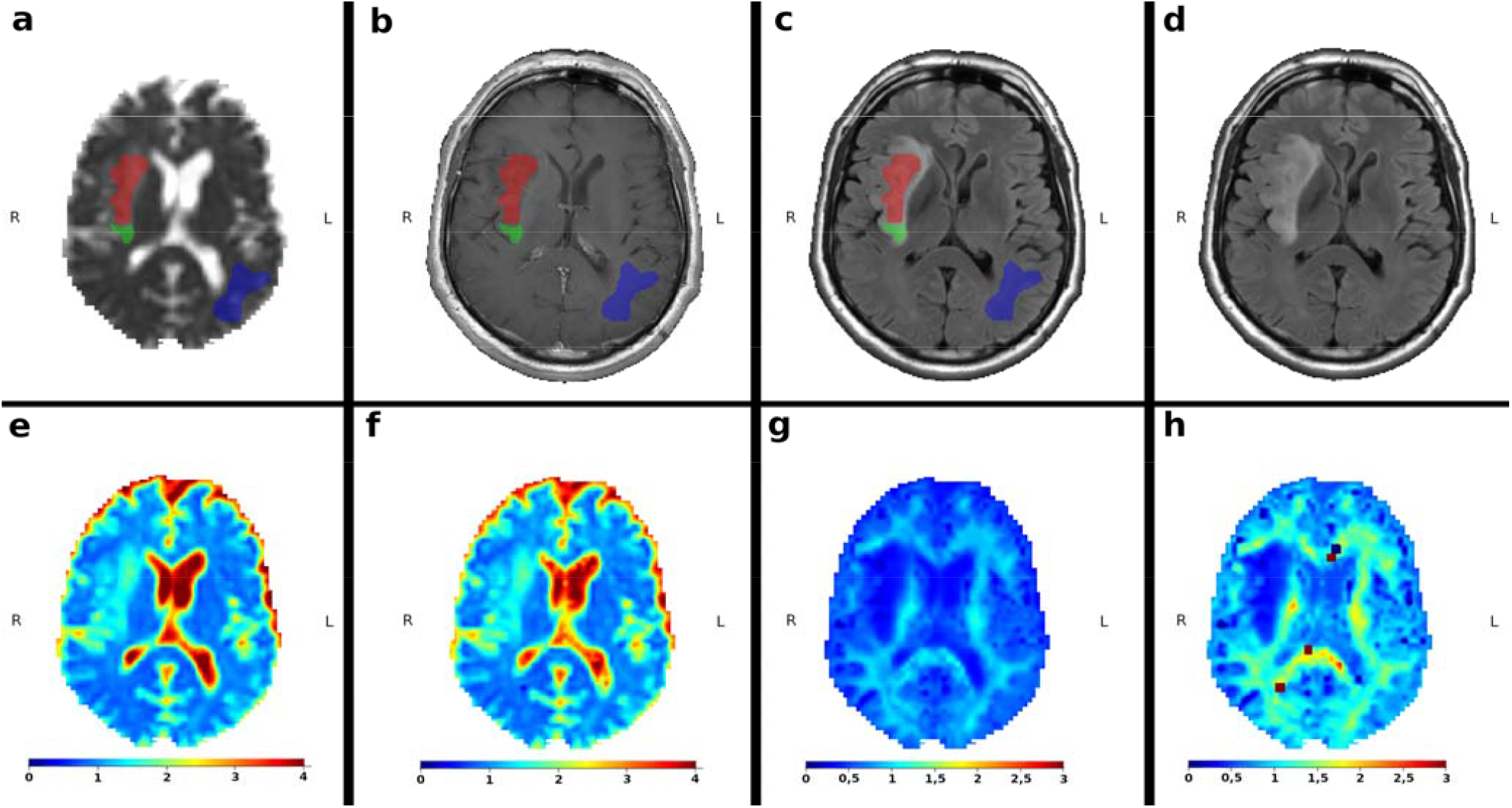
Example of low grade gliomas with the marked regions of interests: red is the tumour, green is the oedema, and blue is the NAWM. All maps are already aligned and interpolated to T_1_-weighted image. **a)** mean diffusivity map with masks; **b)** T_1_-weighted image with contrast agent; **c)** T_2_-weighted image with contrast agent; **d)** T_2_-weighted image without contrast agent; **e)** mean diffusivity map obtained from the conventional kurtosis approach; **f)** mean diffusivity map obtained from the generalised kurtosis approach; **g)** mean kurtosis map obtained from the conventional kurtosis approach; **h)** mean kurtosis map obtained from the generalised kurtosis approach

**Figure 2.**
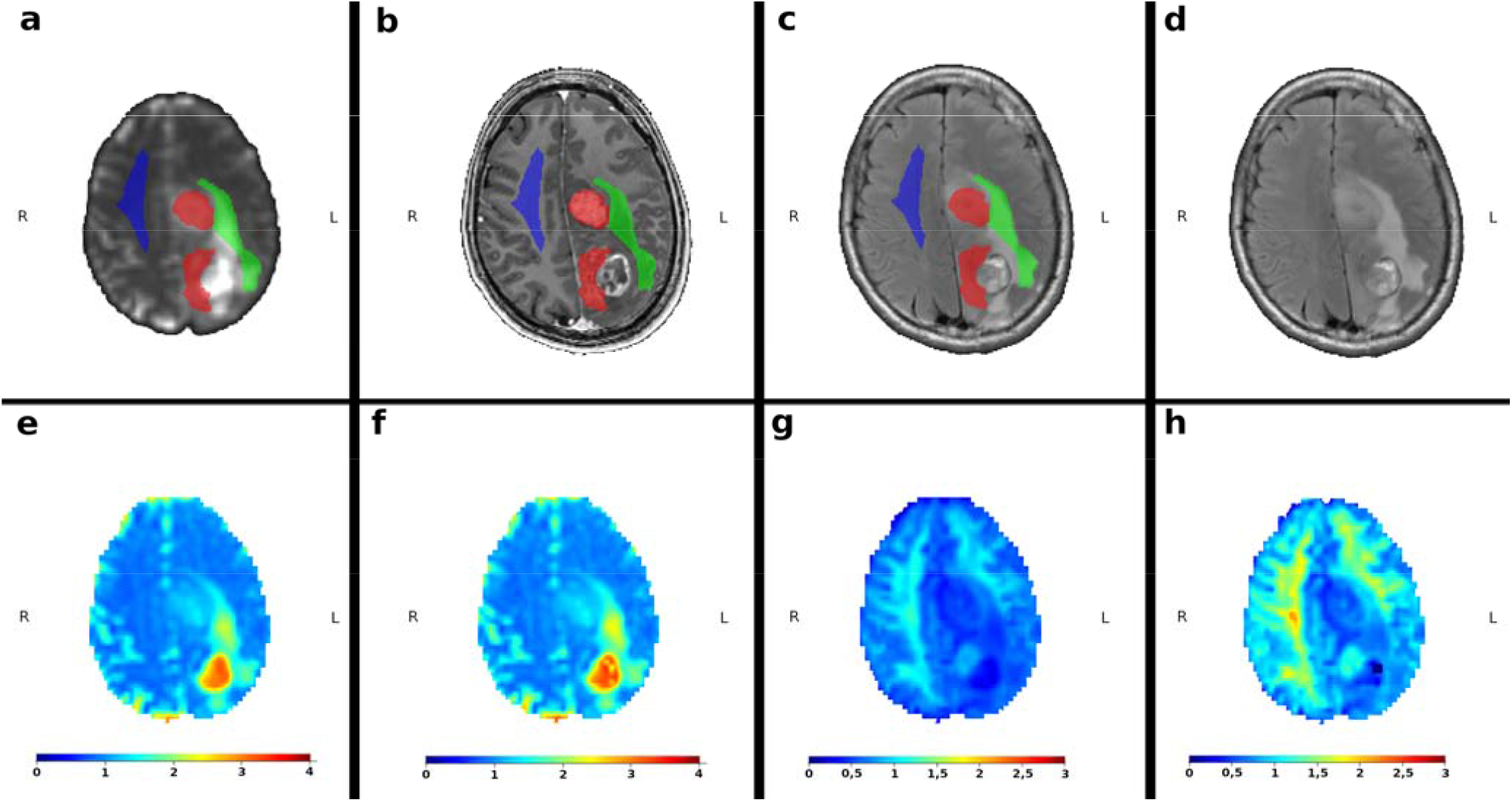
Example of high grade gliomas with the marked regions of interests: red is the tumour, green is the oedema, and blue is the NAWM. All maps are already aligned and interpolated to T_1_-weighted image. **a)** mean diffusivity map with masks; **b)** T_1_-weighted image with contrast agent; **c)** T_2_-weighted image with contrast agent; **d)** T_2_-weighted image without contrast agent; **e)** mean diffusivity map obtained from the conventional kurtosis approach; **f)** mean diffusivity map obtained from the generalised kurtosis approach; **g)** mean kurtosis map obtained from the conventional kurtosis approach; **h)** mean kurtosis map obtained from the generalised kurtosis approach

### Statistical analysis

Statistical analyses were performed using *R* Project (http://r-project.org) and in-house MATLAB scripts. Some voxels among the patient maps contained of corrupted diffusion metrics derived from the DTI and DKI tensors. In order to prevent any statistical bias in analysis, we excluded such voxels from consideration using inequalities: MK < 0 and MK > 3, for both CK and GK approaches. Mean values and standard deviations of all diffusion metrics were calculated from the tumour, oedema and NAWM regions. These values were used for the low- and high-grade glioma differentiation and assessed with a Mann-Whitney-Wilcox U-test with statistical significance reported at the 0.05 level. Effect sizes were calculated with Cohen’s *d* in order to estimate the magnitude of the differences between the conditions/groups. Correlations between the conventional and generalised DKI metrics have been estimated using a linear regression model: *y* = *k*_0_ + *k*_1_•*x*, and package *stat* from *R* Project. Receiver operation characteristic (ROC) analysis was performed using package *pROC* in order to compute the area under curve (AUC) for estimation of the diffusion model performance for tumour differentiation. The squared Pearson correlation coefficients were estimated using the package *stat* from *R* Project.

## Results

Figure 1 and 2 are visual examples of manual masks of three regions, i.e. tumour, oedema, and NAWM together with examples of MD and MK maps for the low- and high-grade glioma patients, estimated by the CK and GK approaches. All maps are presented in T_1_-weighted space with a spatial interpolation of diffusion maps. Briefly, we can see that the visual quality of MD maps between CK and GK approaches is very similar. In turn, as already was shown in [24], the tissue contrast for DKI maps, e.g. MK, is higher for GK approach. Squared outlier voxels both in Figs. 1 and 2 are the result of fitting problems due to noisy diffusion signal. These voxels have been excluded from the analysis.

Figures 3 and 4 present the correlations of diffusion metrics obtained by CK and GK signal expansions. Figure 3 shows DTI metrics only, i.e. FA, MD, AD, and RD with corresponding R-squared values which in the case of linear correlations coincide with the squared Pearson correlation coefficients (see Table 1). In Fig. 4 we present DKI metrics only, i.e. KA, MK, AK, and RK with corresponding R-squared values (see Tab. 1). The scatter plots were estimated voxelwisely for the tumour masks only from low- and high-grade gliomas, respectively. In short, linear correlations between CK and GK estimations are higher for the high-grade glioma in contrast to the low grade glioma. This effect is present for all DTI metrics. In the case of DKI metrics, it applies only for KA and AK metrics. Interestingly, MK and RK metrics have two linearly dependent subgroups of voxels (see, for example, Fig. 4: LGG), which merge as glioma malignancy increases (see, Fig. 4: HGG).

**Table 1.** The results for linear correlations between diffusion metrics obtained from CK and GK approaches (see Figs. 3 and 4). The linear model is *y* = *b*_0_ + *b*_1_·*x*, R-square is the coefficient of determination also coinciding with the squared Pearson correlation coefficient. LGG – low grade glioma, HGG – high grade glioma.

| **LGG** | **FA** | **MD** | **AD** | **RD** | **KA** | **MK** | **AK** | **RK** |
| --- | --- | --- | --- | --- | --- | --- | --- | --- |
| intercept $b_0$ | 0.053 | -0.017 | 0.091 | 0.025 | 0.19 | -0.12 | 0.0088 | -0.15 |
| slope $b_1$ | 0.73 | 1.1 | 1.0 | 1.0 | 0.88 | 1.7 | 1.7 | 1.6 |
| R-square | 0.693 | 0.981 | 0.906 | 0.97 | 0.463 | 0.944 | 0.777 | 0.942 |
| **HGG** |  |  |  |  |  |  |  |  |
| intercept $b_0$ | 0.028 | 0.022 | 0.054 | 0.028 | 0.14 | -0.1 | -0.018 | -0.13 |
| slope $b_1$ | 0.85 | 1.0 | 1.0 | 1.0 | 0.93 | 1.6 | 1.7 | 1.6 |
| R-square | 0.814 | 0.977 | 0.953 | 0.97 | 0.756 | 0.889 | 0.804 | 0.855 |

**Figure 3.**
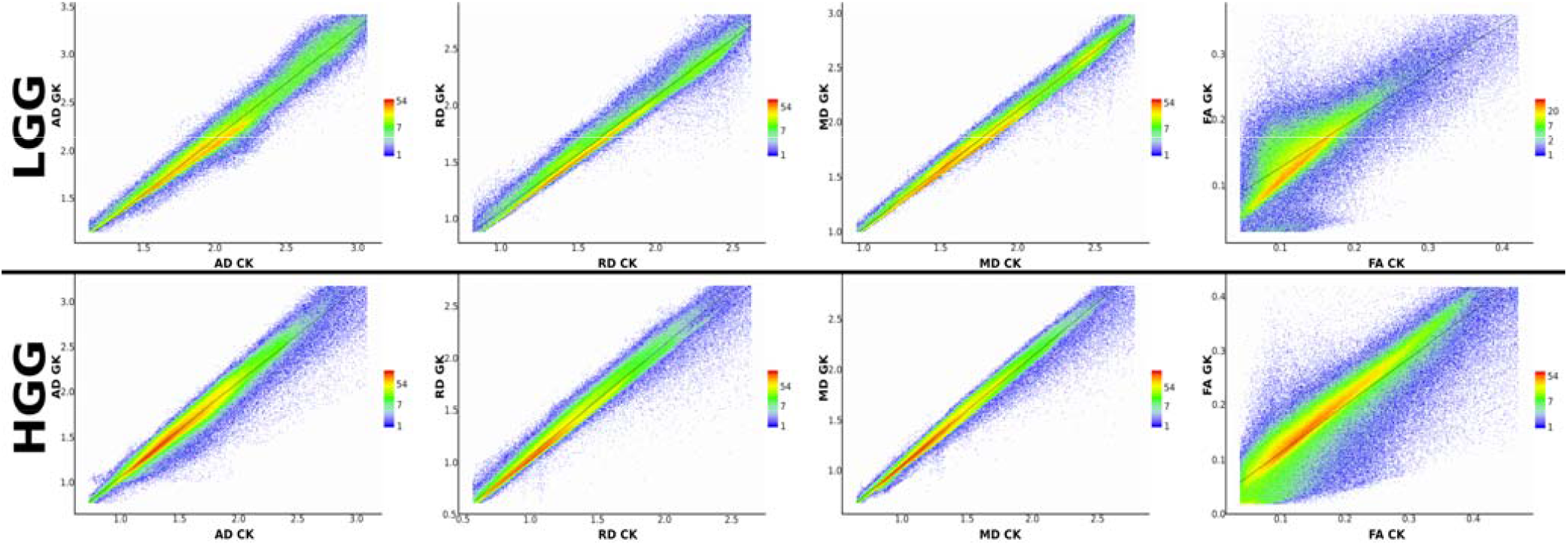
Voxelwise correlations of the diffusion tensor metrics between the conventional and generalised kurtosis approaches. The scalar metrics belong to the tumour masks in low and high grade gliomas for all patients. The columns of plot are axial, radial, and mean diffusivities and fractional anisotropy, respectively

**Figure 4.**
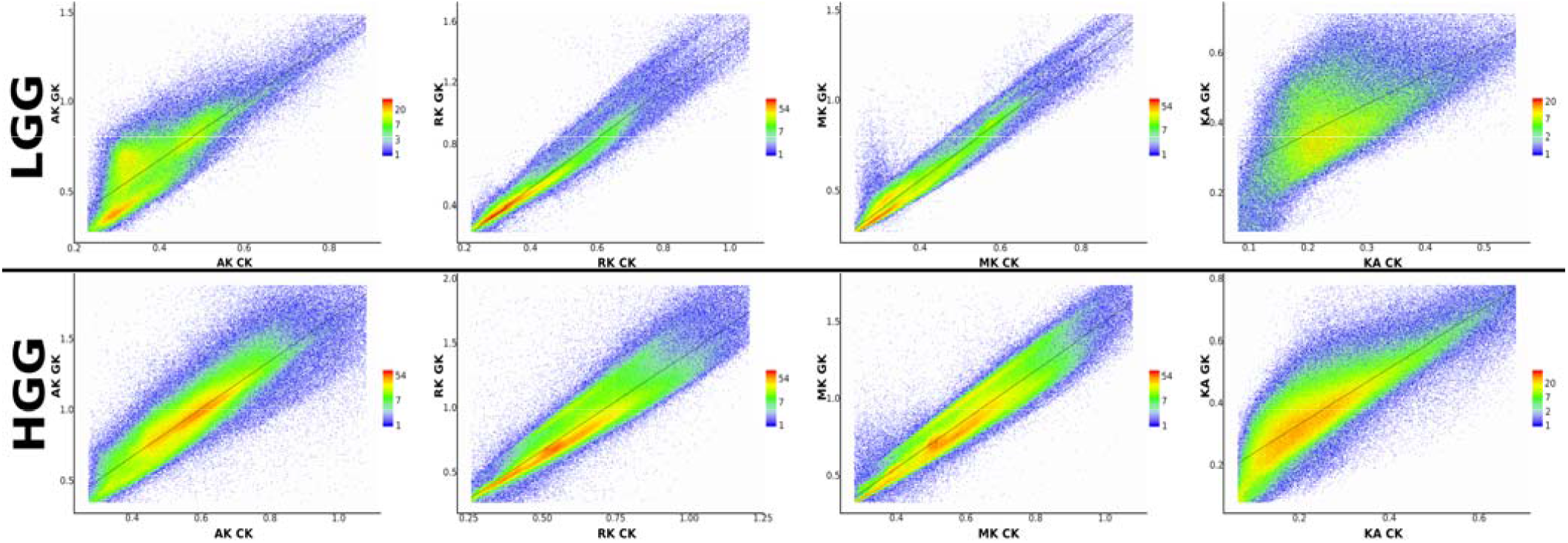
Voxelwise correlations of the diffusion kurtosis metrics between the conventional and generalised kurtosis approaches. The scalar metrics belong to the tumour masks in low and high grade gliomas for all patients. The columns of plot are axial, radial, and mean kurtosis and fractional anisotropy of kurtosis, respectively

In Figure 5 we present boxplots of the mask-averaged diffusion metrics estimated from CK and GK approaches. The averages were performed for each ROI such as tumour, oedema, and NAWM for each patient. The boxplots are structured as ROI-wise and glioma grades pairs. For each pair of low- and high-grade gliomas, we performed the Mann-Whitney-Wilcox U-test in order to check for significant (p < 0.05) differences between glioma grades in all regions including tumour, oedema and NAWM. There were no significant differences between low- and high-grade glioma diffusion metrics for oedema, and there was difference between GK metrics of RD for NAWM. However, for tumour regions, both CK and GK metrics except for KA and FA, we revealed a significant difference for low- and high-grade gliomas (marked by asterisk in Fig. 5). In the case of FA metrics, GK approach did not reveal significant differences between glioma grades. The effect sizes for the diffusion metric differences detected by both CK and GK approaches are summarised in Table 2. For DTI metrics, the effect size is large for both kurtosis approaches, while for DKI metrics the differentiation possesses very large effect size except for GK-derived RK metric. We also performed a pair comparison between CK and GK metrics marked by the cyan background colour for significant differences (p < 0.05) between the CK-GK metrics. Interestingly, for all DKI metrics and ROIs, there were significant difference between CK and GK metrics. However, for DTI metrics, the significant difference was detected only for NAWM regions for FA, MD, and RD metrics.

**Table 2.** The effect size of significant differences between low and high grade gliomas detected by CK and GK derived diffusion metrics (see Fig. 5). The effect size is estimated using the Cohen’s *d*.

| **Effect size** | **FA** | **MD** | **AD** | **RD** | **KA** | **MK** | **AK** | **RK** |
| --- | --- | --- | --- | --- | --- | --- | --- | --- |
| **CK** | 0.99 | 1.21 | 1.11 | 1.23 | 0.11 | 1.56 | 1.47 | 1.52 |
| **GK** | 0.65 | 1.15 | 1.13 | 1.15 | 0.30 | 1.50 | 1.35 | 0.24 |

**Figure 5.**
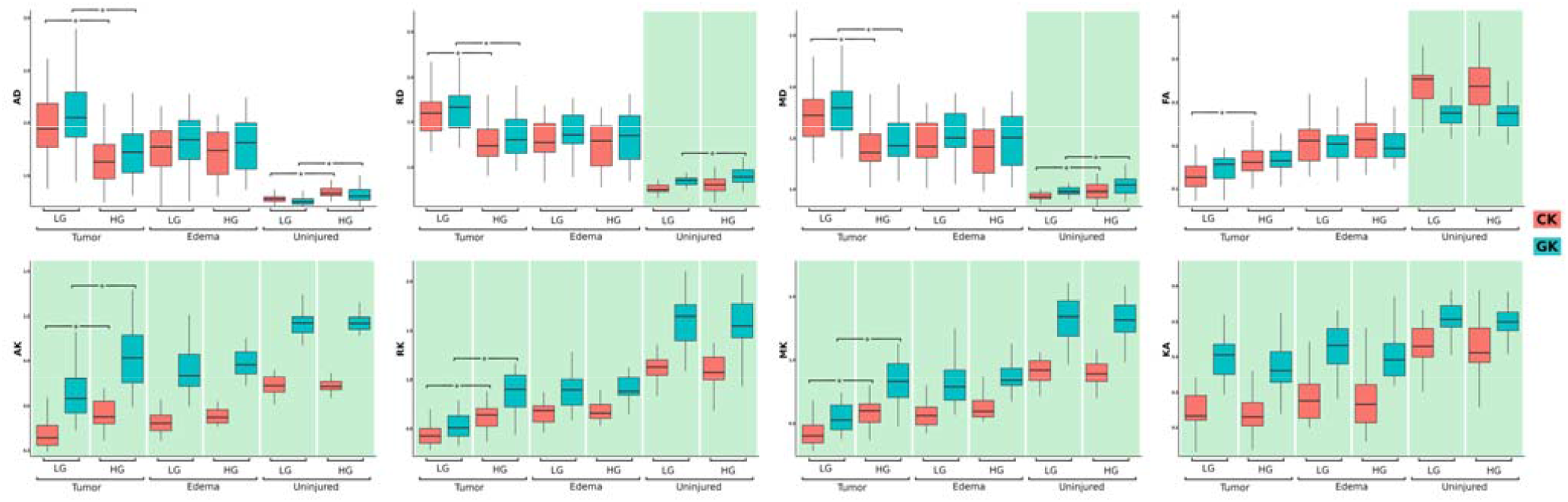
The boxplots of averaged diffusion tensor and kurtosis metrics in different regions: tumour, oedema, and normally appeared white matter. The star-marked brackets exhibit the significant (p < 0.05) difference between low and high grade gliomas for the diffusion metrics. The green background colour emphasise the significant difference between diffusion metrics for the conventional and generalised approaches

It is of vital importance to perform a comparison between CK and GK approaches in terms of their sensitivity and specificity. These results are presented in Figure 6. Both approaches demonstrated high rate of sensitivity and specificity for low and high glioma discrimination with negligible difference between CK and GK approaches. The estimated sensitivity, specificity, cutoff and area under curves (AUC) are summarised in Table 3.

**Table 3.** Differentiation performance for diffusion metrics estimated by CK and GK approaches. AUC of ROC (see Fig. 6), cutoff, senstivity and specificity values are presented.

| **CK** | **FA** | **MD** | **AD** | **RD** | **KA** | **MK** | **AK** | **RK** |
| --- | --- | --- | --- | --- | --- | --- | --- | --- |
| **AUC** | 0.77 | 0.79 | 0.78 | 0.80 | 0.53 | 0.87 | 0.86 | 0.86 |
| **cutoff** | 0.15 | 1.50 | 1.72 | 1.39 | 0.23 | 0.53 | 0.41 | 0.54 |
| **sensitivity** | 0.74 | 0.71 | 0.71 | 0.71 | 0.48 | 0.74 | 0.93 | 0.74 |
| **specificity** | 0.74 | 0.84 | 0.84 | 0.84 | 0.68 | 0.89 | 0.68 | 0.90 |
| **GK** |  |  |  |  |  |  |  |  |
| **AUC** | 0.65 | 0.78 | 0.77 | 0.78 | 0.62 | 0.86 | 0.83 | 0.88 |
| **cutoff** | 0.17 | 1.71 | 1.80 | 1.59 | 0.39 | 0.73 | 0.81 | 0.71 |
| **sensitivity** | 0.48 | 0.80 | 0.71 | 0.81 | 0.68 | 0.74 | 0.71 | 0.77 |
| **specificity** | 0.74 | 0.68 | 0.84 | 0.68 | 0.68 | 0.89 | 0.89 | 0.89 |

**Figure 6.**
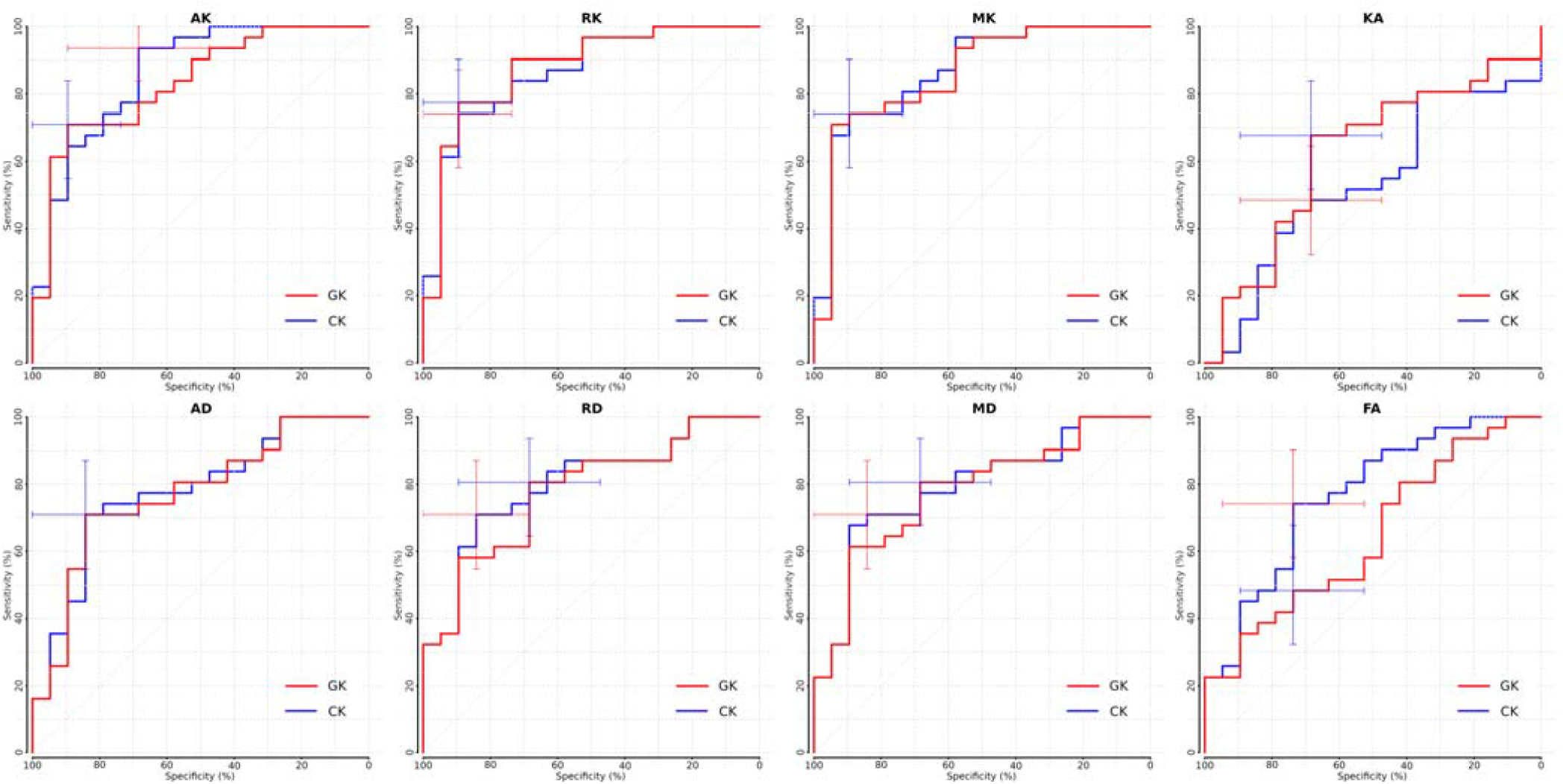
Reciever operating curves (ROC) for the conventional (CK) and generalised kurtosis (GK) approaches for differentiation of low and high grade gliomas. The error-bars exhibits the detected sensitvity and specificty of both methods

## Discussion

In the present study we estimated two diffusion imaging approaches based on the conventional and generalised kurtosis expansions. The scalar metrics based on GK approach demonstrated higher linear correlations with CK derived metrics for high-grade glioma and comparable sensitivity and specificity in glioma differentiation between low- and high-grade gliomas. In turn, the scalar diffusion metrics based on GK approach exhibited brighter tumour and peritumoural oedema contrasts compared to healthy tissue allowing clinicians to better visualise tumour spreading in a healthy brain tissue environment.

The tumour grade differentiation constitutes a challenge for many imaging techniques such as MRI, CT, or PET. This is due to the need of a quantitative parametrisation enabling both the assessment of damage of healthy brain tissue by aggressive growth of tumour and the degree of the malignancy of this process. This is particularly complicated due to multifarous forms of tumour manifestation, including cell swelling, microvasculature proliferation, tumour heterogeneity, presence of necrosis. Thus, the diffusion MR imaging grants two levels of tumour assessment, that is easy-to-interpret scalar maps and sensitivity of these maps to the micrometer scale of tissue changes. Therefore, diffusion kurtosis imaging offers an excellent tool for probing brain tumour, enabling estimation of the qualitative and quantitative differences between different glioma grades and their dynamics.

The difference between the conventional and generalised kurtosis estimations is defined by a fraction multiplyer α and a quadratic kurtosis term in Eq. (2). It is interesting that dispersion in the distribution between DTI metrics (see Fig. 3) is quite low and does not depend on glioma grade. However, in the case of kurtosis scalar maps (see Fig. 4), in particular for MK and RK, there is a stronger value spreading in low glioma metric correlations. Such kind of distribution behaviour can indicate two different types of tissue organisation present at early stages of the glioma tumour: resting healthy brain tissue and highly heterogeneous tumour. As glioma malignancy increases, the heterogeneous tumour tissue becomes dominant due to aggressive growth which leads to higher correlations between CK and GK metrics, i.e. quadratic kurtosis term might drive this effect. We see that GK-derived MK and RK metrics are more sensitive to the tumour heterogeneity in low-grade glioma. Therefore, a variation in data dispersion between CK and GK metrics, in particular, in kurtosis metrics, might provide us with an additional information about glioma grades. Notably, this information is accessible only for voxelwise assessment, where typical glioma grade differentiation is performed for region-averaged diffusion metrics.

This hypothesis is supported by significant differences between CK and GK metrics for kurtosis metrics in contrast to the DTI ones (see Fig. 5). For kurtosis metrics, the significant differences between CK and GK derived values were found in all regions: tumour, oedema, and NAWM. On the other hand, DTI metrics demonstrated significant differences only for normally appearing brain tissue, likely due to the different regions of the brain chosen as NAWM and strong age dependence of the patients. Thus, the quadratic kurtosis term strongly influences the kurtosis scalar metrics while it does not affect the DTI ones.

It is quite important to estimate general brain changes associated with tumour grades in accordance with healthy tissue. All DTI metrics (FA, MD, AD, and RD) demonstrated their potential as a glioma grade biomarker, except for FA maps derived from GK (see Tabs. 2 and 3). The values of MD, AD, and RD show lower diffusivities in high grade glioma for both CK and GK approaches. In turn, diffusion metrics for both low- and high-grade gliomas demonstrated higher diffusivities in contrast to the NAWM values. This effect might be related to damage of healthy tissue as extra-cellular space increases accompanied by simultaneous microvascularisation process in the tumour tissue. Interestingly, low-grade glioma tissue retains higher diffusivities in contrast to high grade one, likely due to an increased cell proliferation rate in high-grade glioma, i.e. emergence of more cellular barriers limiting the water diffusion.

Kurtosis scalar maps exhibited the same trends as DTI metrics for low and high glioma differentiation and the contrast between the tumour and healthy tissue regions. Notably, KA did not reveal significant differences between low- and high-grade gliomas. Nonetheless, MK, AK, and RK metrics demonstrated lower kurtosis values in low-grade glioma in contrast to both high-grade glioma and healthy tissue (see Fig. 5).

We did not find significant differences for oedema regions for neither low nor high grade gliomas. At the same time, oedema metrics exhibited close range of kurtosis and DTI values with high grade glioma, suggesting that the peritumoural tissue organisation is more complex than free water diffusion. This effect may be caused by the compression of the surrounding cells and resting microstructure of healthy tissue. However, this hypothesis should be verified and examined with different types of glioma spreading processes such as the level of cell infiltration that demands high resolution diffusion measurements.

As a limitation of the study we should note not an optimal choice of adjusting variable α. A more accurate model fitted to the tumour microstructure [33] might increase feasibility of GK approach and its performance in the case of glioma grading. Addtionally, advanced diffusion techniques including fast kurtosis [34], isotropic diffusion weighting [35], [36], [37] and multidimensional diffusion imaging [38], [39] might help clinicians to perform a robust, fast and non-invasive glioma differentiation. We plan to implement and apply these approaches for research and patiant treatment in the future.

In conclusion, the generalised diffusion kurtosis imaging presents an additional source of information enabling differentiation of low- and high-grade gliomas at the same level as the conventional DKI. The GK approach exhibited higher tissue contrast and, thus, offers more sensitive scalar maps to glioma tissue heterogeneity.

## Data Availability

data are not shared

## Declaration

### Ethical approval

All procedures performed in study involving human participants were in accordance with the ethical standards of the institutional and/or national research committee and with the 1964 Helsinki declaration and its later amendments or comparable ethical standards.

### Informed consent

Informed consent was obtained from all individual participants included in the study.

